# Hemodialysis Prescribing Patterns of Hospital & Satellite Centres: An Institution-Wide Observational Study

**DOI:** 10.64898/2026.04.20.26351284

**Authors:** Sarah Melville, Martin G. MacKinnon, Jacob Michaud

## Abstract

**Background:** Life-sustaining hemodialysis (HD) is onerous for patients, especially those with multiple co-morbidities and advanced age. A standard HD prescription is 720 minutes per week. Alternative HD regiments have been proposed in attempt to maintain quality of life (QOL). Studies are needed to investigate the efficacy and safety of less frequent HD prescriptions in this population. This is an institution-wide observational study in New Brunswick, Canada to compare HD prescriptions and the impact on QOL and mortality.

**Objective:** The purpose of this study is to assess the current HD prescribing practices at a provincial healthcare institution in relation to patient QOL.

**Design:** Prospective Observational Study.

**Setting:** Single centre hospital and satellite hemodialysis units.

**Patients:** Voluntarily consented patients undergoing in-centre hemodialysis treatment.

**Measurements:** Observational clinical data was collected for each study participant from their hospital and dialysis electronic medical records. The KDQOL-36^TM^ questionnaire was used to assess patient-reported quality of life at the time of consent.

**Methods:** Adults undergoing in-centre or satellite site HD for at least 3 months were eligible to participate. Consenting patient participants were grouped by HD prescription whether they were prescribed 720 minutes or more per week or less than 720 minutes per week. All participants completed the KDQOL-36 ^TM^ questionnaire to estimate QOL and groups were compared using the Mann-Whitney U statistical test. Emergency department visits, hospitalizations, and mortality were analyzed using a negative binomial regression or a logistic regression.

**Results:** We enrolled 140 patient participants; 41 were undergoing less than 720 minutes per week of HD and 99 were undergoing 720 minutes or more of HD per week. Patients who were undergoing less than 720 minutes per week of HD were older [Median (IQR): 76 (72- 81) yrs. vs. 64 (55 - 75) yrs.; p < 0.001], had higher median (IQR) QOL scores on the ‘Symptoms/ Problems List’ scale on the KDQOL-36 ^TM^ questionnaire [79.2 (70.8 – 88.5 vs. 70.8 (62.5 – 81.3); p = 0.0022], and were less likely to present to the emergency department (incident rate ratio 0.52, 95% confidence interval [CI] 0.33-0.81). Mortality was similar between groups, even when adjusted for age and comorbidity score (odds ratio 1.62, 95% CI 0.59-4.49).

**Limitations:** Patient participant enrollment was limited by the single centre nature of this study. As this was an observational study, we did not account for how long the patients had been prescribed less than 720 minutes of hemodialysis. We did not include a frailty assessment of the study participants. A higher number of study participants may have identified significant trends in mortality.

**Conclusions:** The results of this study show that patients undergoing less than 720 minutes of weekly HD had a higher QOL score for the ‘KDQOL-36 ^TM^ ‘Symptoms/ Problems List’ scale, were less frequently in the emergency department and were not more likely to die than patients undergoing 720 minutes or more of weekly HD. Further studies are required to assess the feasibility and safety of a conservative model of HD prescribing to improve QOL of patients with palliative care treatment goals.

## Introduction

Hemodialysis (HD), a life-sustaining therapy for people living with kidney failure, was once offered only to patients deemed suitable by their nephrologists; often younger patients with minimal comorbidities.^1^ As universal access to HD has become entrenched in Canada, HD patients are older, have multiple comorbidities, and present in higher numbers than ever before.^2,3^ When patients with kidney failure meet dialysis initiation criteria (and in-centre HD rather than other forms of dialysis is preferred by the patient and/or deemed most appropriate), HD is usually offered to patients in three weekly 4-hour sessions (720 minutes per week) to achieve predetermined HD quality measures.^4,5^ While treatment with HD is life-sustaining, it is associated with multiple potential complications such as vascular access site infections, loss of residual kidney function and dialysis-dependence as well as increased incidence of major adverse cardiac events and nutritional issues; all of which may contribute to significant morbidity and mortality.^6–8^ In fact, adjusted mortality rate of patients who are receiving dialysis is 10- to 15-fold higher than patients who are not receiving dialysis and mean survival among patients receiving HD is only 47 months after initiation.^9^ For patients who decide to discontinue dialysis, the mean survival time is between 8-10 days.^10^

Patients treated with HD can also experience a high burden of physical and psychological symptoms related to HD treatments which negatively impact mood and quality of life.^11–15^ Considering the increasing fragility of HD patients, the high mortality rates associated with HD, and the symptom burden, there is a growing need for palliative models in dialysis care to optimize quality of life.^3,13,16,17^ Previous publications have outlined strategies used to reorient dialysis care from achieving life-sustaining standards of care to focusing on patient-centred metrics and adequate control of symptoms. This personalized or individualized approach to HD treatment includes strategies such as: (1) reducing dialysis sessions length and/or dialysis frequency, (2) less emphasis on dietary restrictions, (3) less frequent bloodwork to monitor chronic kidney disease-related mineral and bone disorders (CKD-MBD) or anaemia, (4) permissive hypertension, (5) pain and dialysis-related symptom management, and (6) use of central venous catheters (CVC) instead of other preferred vascular access methods (such as arteriovenous fistula or grafts).^3,13^ We have informally observed a trend towards palliative prescription patterns in New Brunswick, specifically in the reduction of HD session time and/or frequency to less than 720 minutes per week for some older patients, irrespective of small molecule clearance or other measures of assessing the quality of dialysis. There is an evolving practice that the standard HD prescription of 4-hour sessions, thrice weekly may not be suitable for every patient undergoing HD. For example, prescribed session duration and frequency less than 720 minutes per week based on residual kidney function has been used in patients initiating HD in a practice known as ‘incremental dialysis.’^18^ For patients nearing end-of-life, increasing the HD prescription may not align with palliative care treatment goals and patient preferences for quality of life such as spending more time at home and with family.

The purpose of this prospective observational study is to quantitatively describe the current HD prescribing practices at our central HD site and four satellite HD sites in New Brunswick in the context of patient-reported quality of life, emergency department visits, hospitalizations, medication burdens, laboratory values, and mortality. The results of this study could qualify the impact of quality of life and mortality of prescribing reduced-dose HD as a strategy to mitigate dialysis burden for older patients with palliative care treatment goals.

## Methods

### Patient population

This investigator-initiated observational study was approved by the Horizon Health Network (HHN) Research Ethics Board (RS#: 2023-3305), and conducted between May 31, 2024 and July 27, 2025. Eligible patients were recruited from the HHN Saint John Regional Hospital and the four satellite HD sites. All voluntary study patient participants provided written informed consent. The study inclusion criteria were: receiving maintenance HD treatment within the jurisdiction of the health authority with a HD vintage of at least 12 weeks, age 19 years or older, residence in New Brunswick for at least one year, not currently active on the transplant list, not admitted to the hospital within the past 4 weeks, and willing to voluntarily participate and sign the study informed consent form.

### Data collection

Data was collected for each study participant from their hospital and dialysis electronic medical records and included: HD prescription, average HD session time for 4 weeks from the date of consent, age, sex, height, prescribed weight, body mass index (BMI), HD vintage, reason for a HD prescription, type of vascular access, total number of medications, number of medications per week, and the assessment of any of 11 diagnosed comorbidities [atherosclerotic heart disease, congestive heart failure, cerebrovascular accident (CVA)/ transient ischemic attack (TIA), peripheral vascular disease, other cardiac diseases (i.e. pericarditis, endocarditis, myocarditis, heart transplant, heart valve replacement, cardiac devices), chronic obstructive pulmonary disease (COPD), gastrointestinal bleeding, liver disease, dysrhythmia, cancer, and diabetes] as per the comorbidity index developed by Liu et al. which has been previously shown to correlate with mortality in dialysis patients.^19^ A province-wide electronic database review was done to collect the number of emergency department visits, the number of days hospitalized, and the laboratory values of serum albumin, phosphate, calcium, and parathyroid hormone. We assessed medication burden by counting the total medications per participant and counting the number of pills (oral) and injections (parenteral) per week per participant, as previously described.^20^ The median ultrafiltration rate (UFR) and the mean urea clearance estimated by Kt/V Diascan (Artis Physio System, Baxter Healthcare Corporation, Deerfield, IL, USA) were collected from each of the HD sessions for the 4 weeks from the date of consent. We also collected mortality data of the consented patient participants up to one year after the date of consent.

Each participant was asked to complete the Kidney Disease Quality of Life (KDQOL)-36^TM^ questionnaire^21^ in the language of their choice (English or French) after voluntarily consenting to participate in the study. The KDQOL-36 ^TM^ questionnaire is a commonly used and validated instrument to assess patient-reported quality of life of people living with chronic kidney disease who are undergoing a dialysis treatment modality. This quantitative questionnaire comprises of 36 multiple-choice questions and the responses are scored on a 0 to 100 range using a pre-designed scoring template. Each question corresponds to one of five scales: Symptoms & Problems List, Burdens of Kidney Disease, Effects of Kidney Disease, Short Form (SF)-12 Physical Health Composite, and SF-12 Mental Health Composite. The higher the score on each respective scale, then the better the reported quality of life.^22^

### Mitigating Bias

To minimize bias, all patients receiving maintenance HD within the health network who met the predefined study inclusion criteria were invited to volunteer to participate in the study which reduced the risk of selective recruitment. Clinical data were obtained from electronic medical records and provincial databases to ensure objective outcome ascertainment. Mortality analyses were adjusted for age and comorbidity to account for baseline differences between groups. For the KDQOL-36^TM^ questionnaire to assess patient-reported quality of life, the participants completed the questionnaire independently without interpretation of the questions by the research team and in accordance with the KDQOL-36^TM^ questionnaire user’s manual instructions.^22^

### Statistical Analysis

Patients were grouped according to their prescribed HD frequency and session duration in two groups: patients who were prescribed less than 720 minutes of HD per week and patients who were prescribed 720 minutes or more of HD per week. We used descriptive statistical methods, scatter plots, and multivariate analyses and a forest plot to compare the participants based on their HD prescription as of the date of consent. Emergency department visit and hospitalization data were analyzed with negative binomial regression due to over-dispersed, right-skewed count distributions with a substantial proportion of zero events, and mortality data were analyzed with logistic regression adjusted for age and comorbidity scores as continuous variables. We used *Stata* version 18 (StataCorp LLC, College Station, TX, USA) for these analyses and *GraphPad Prism* version 10.5.0 for Windows (GraphPad Software, LLC, Boston, MA USA, www.graphpad.com) for the data visualizations.

This study is reported in accordance with the *Strengthening the Reporting of Observational Studies in Epidemiology* (STROBE) reporting guidelines (STROBE Checklist; Supplementary 1).^23^

## Results

We screened 150 potentially eligible patient participants, of which 142 patient participants voluntarily consented. There were 2 participants who moved out of province who were withdrawn from the data analysis due to lack of access to medical records for the study data collection, therefore data was collected and analyzed for a total of 140 study patient participants. The STROBE flow diagram of the study procedures and data collection is illustrated in Figure 1.

**Figure 1:**
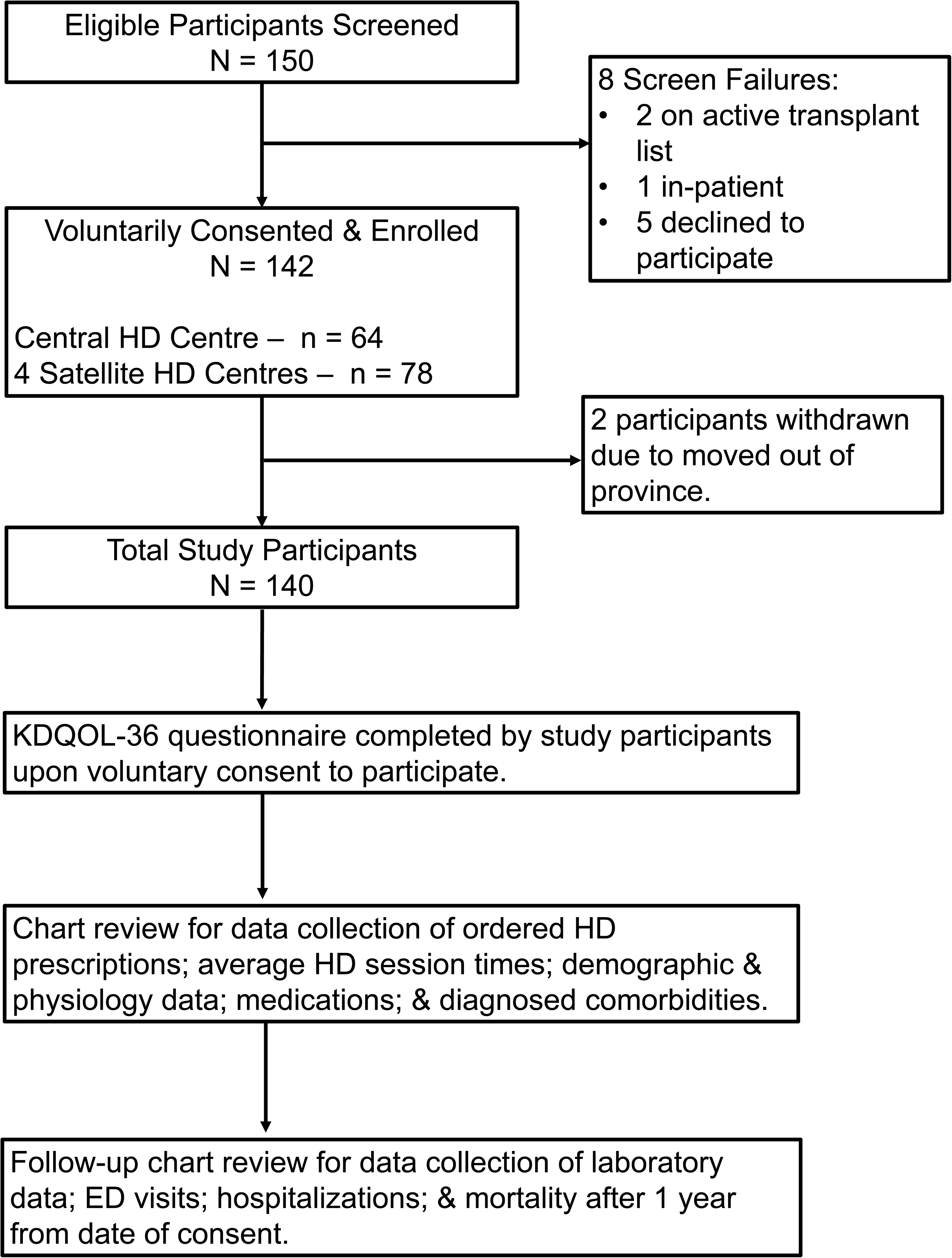
STROBE Diagram of Study Procedures. The STROBE flow diagram of the study procedures of patient participant recruitment and consent, and subsequent data collection.

**Figure 2:**
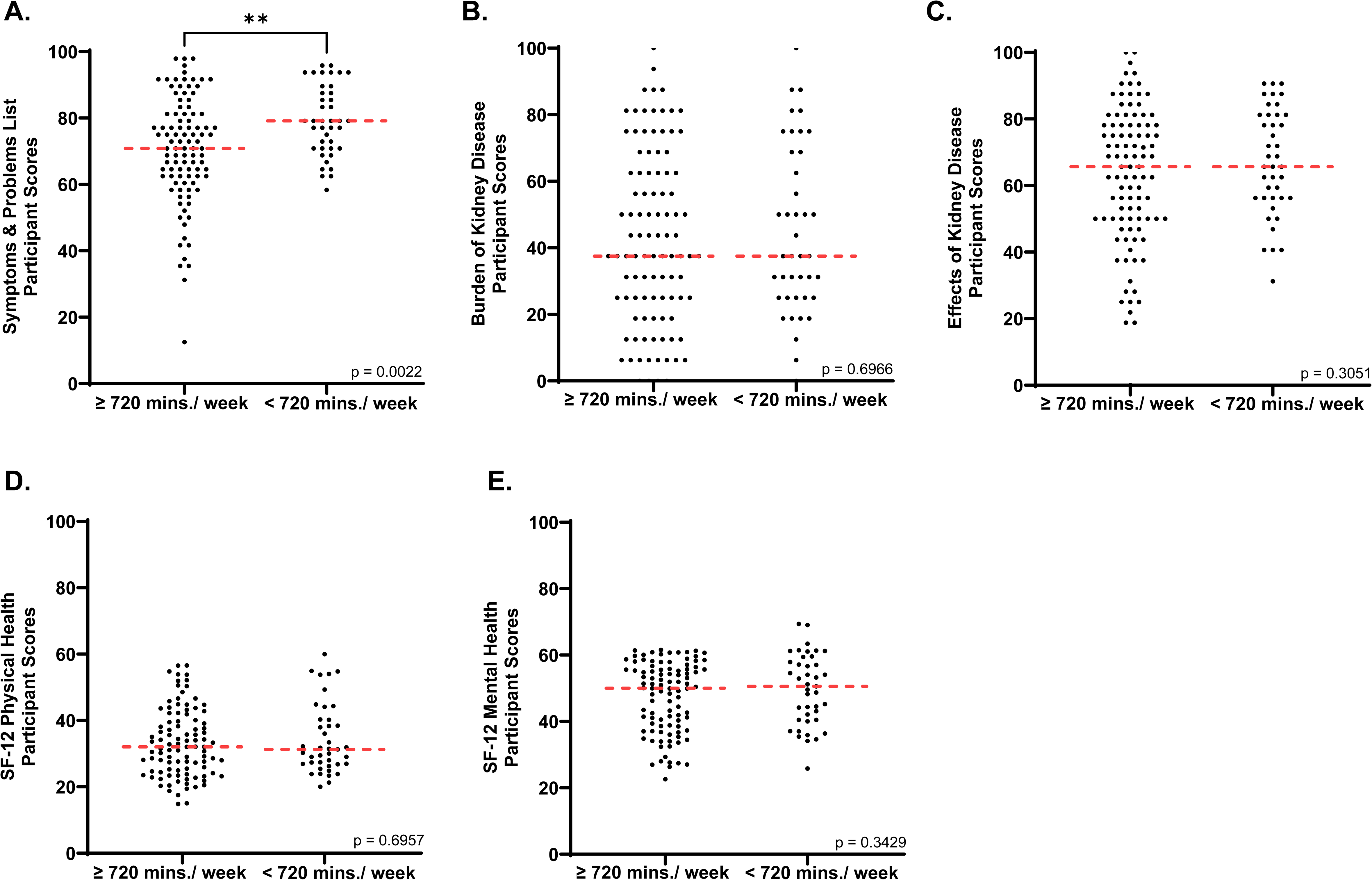
KDQOL-36^TM^ Questionnaire Results. Scatter plots of the KDQOL-36^TM^ questionnaire score results for each of the 5 variables for the study patient participants receiving 720 minutes or more of HD per week or receiving less than 720 minutes of HD per week: (A) Symptoms/ Problems List [Median (IQR) 70.8 (62.5 – 81.3) vs. 79.2 (70.8 – 88.5), p = 0.0022]; (B) Burden of Kidney Disease [Median (IQR) 37.5 (25.0 – 62.5) vs 37.5 (25.0 – 68.8), p = 0.6966]; (C) Effects of Kidney Disease [Median (IQR) 65.6 (50.0 – 78.1) vs. 65.6 (56.3 – 81.3), p = 0.3051]; (D) SF-12 Physical Health Composite [Median (IQR) 32.1 (24.7 – 40.1) vs. 31.3 (26.9 – 40.2), p = 0.6957]; and (E) SF-12 Mental Health Composite [Median (IQR) 50.0 (38.6 – 55.8) vs. 50.6 (40.5 – 58.7), p = 0.3429].

**Figure 3:**
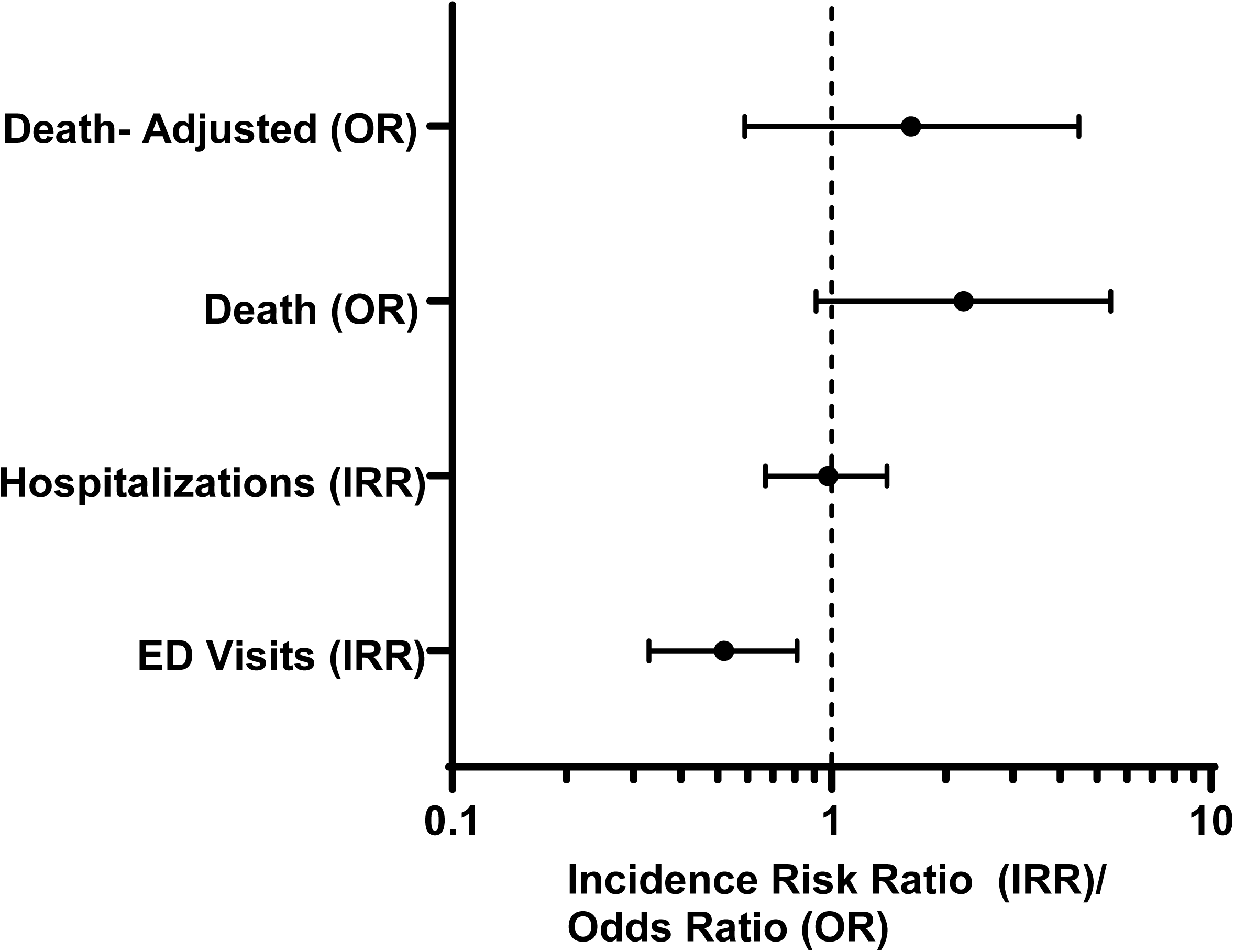
Multivariate Analyses. Forest plot with a logarithmic scale (x-axis) of the multivariate analyses for emergency department visits; hospitalizations; death; and death adjusted for age and comorbidity score.

At the time of consent, 99 participants (70.7%) had a HD prescription of 720 minutes or more per week, and 41 participants (29.3%) had a HD prescription of less than 720 minutes per week. The participants who were prescribed less than 720 minutes of HD were older [Median (IQR): 76 (72 – 81) years vs. 64 (55 – 75) years; p < 0.0001) and had a lower median (IQR) body mass index [23.4 (20.1 – 25.3) vs. 31.8 (24.9 – 35.1); p <0.0001] than the participants who were prescribed 720 minutes or more of HD per week. There were no differences in the median (IQR) total comorbidity scores [3.0 (1.0 – 5.0) vs. 2.0 (1.0 – 5.0); p = 0.6021] nor were there statistical differences between any of the 11 variables used in the comorbidity index score calculation. The central venous catheter was the most common access type in both groups [33 of 41 (80.5%) and 72 of 99 (72.7%) for the less than 720 minutes and the 720 minutes or more of HD, respectively). The complete study participant characteristics are presented in Table 1. .

**Table 1:**
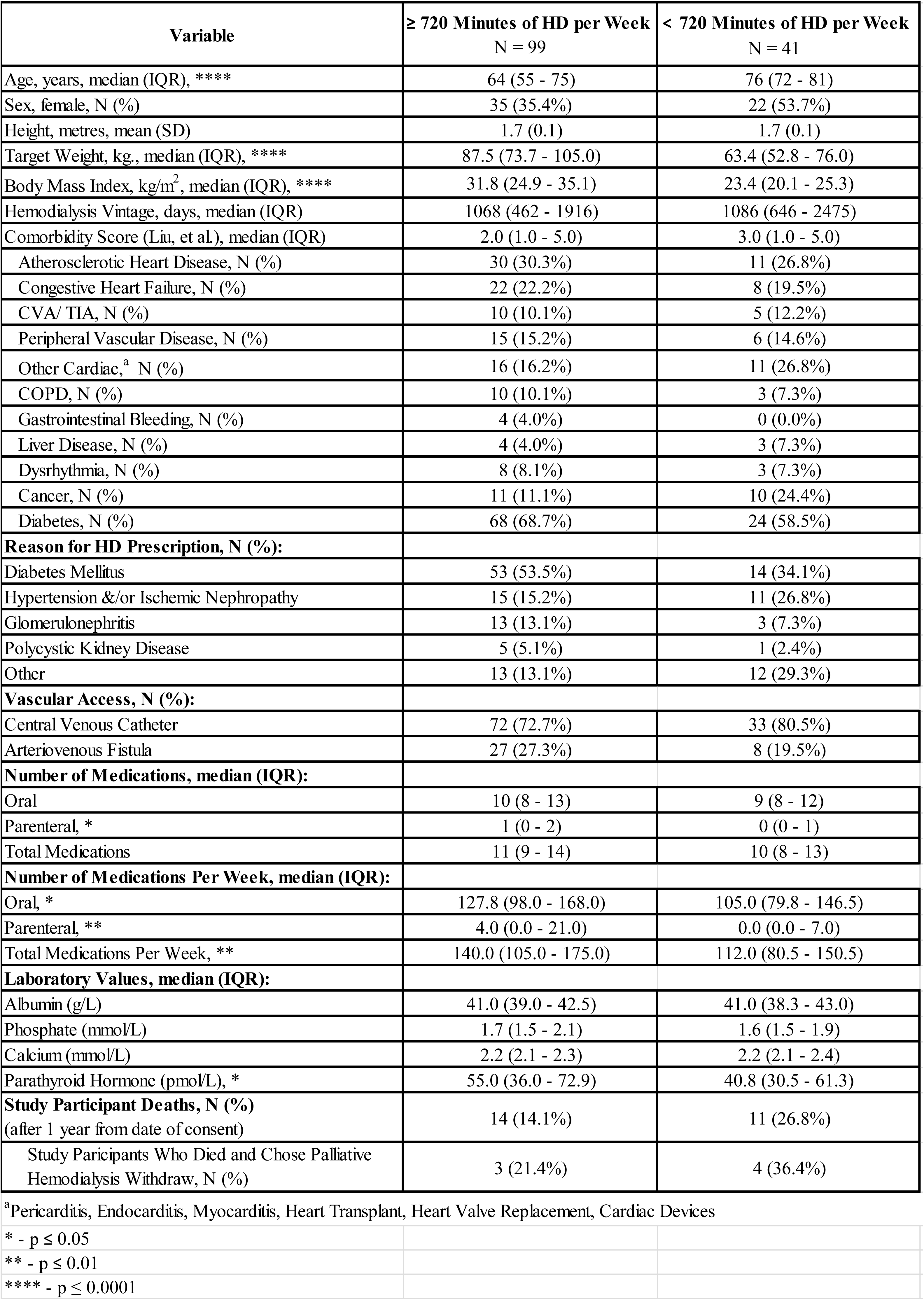
Study Participant Characteristics. The characteristics of the observed study patient participants (N = 140).

Prescribed HD frequencies and durations for each group are shown in Table 2. . In the 720 minutes or more group, most of the participants were prescribed three four-hour sessions per week [93 of 99 (93.9%)], while only 6.1% (6 of 99) of participants were prescribed more than 720 minutes per week. In the less than 720 minutes group, 68.3% (28 of 41) of participants were prescribed thrice weekly HD and 31.7% (13 of 41) of the participants were prescribed twice weekly HD. Of the patient participants who were prescribed twice weekly HD, none of the participants were receiving incremental HD (i.e., a lower prescription when initiating HD based on residual kidney function) as determined by chart review, and there was no plan to increase the HD prescription. The HD prescriptions and the average HD delivered over 4 weeks are shown in Supplementary Figure 1. There was statistically significant less adherence to the HD prescription in the 720 minutes or more group compared with adherence in the less than 720 minutes of HD prescription group [Median (IQR): 97.6% (94.5% - 99.2%) vs. 99.0% (97.6% - 99.9%), p = 0.0311]. Specifically, 26.3% (26 of 99) of patients received less than 95% of their HD prescription versus 22.0% (9 of 41), respectively. (Supplementary Figure 1).

**Table 2:** Hemodialysis (HD) Prescriptions (N = 140) The observed hemodialysis prescriptions of the study patient participants at the time of consent, grouped by 720 minutes or more of hemodialysis per week (A), or less than 720 minutes of hemodialysis per week (B).

**A. 720 Minutes or More of HD per Week**
| **Frequency**<br>(HD sessions per week) | **Duration**<br>(hours per HD session) | **Study Participants**<br>N (%) |
| --- | --- | --- |
| 3 | 4.0 | 93 (66.4) |
| 3 | 4.5 | 2 (1.4) |
| 3 | 5.0 | 1 (0.7) |
| 4 | 4.0 | 2 (1.4) |
| 6 | 3.5 | 1 (0.7) |

**B. Less than 720 Minutes of HD per Week**
| **Frequency**<br>(HD sessions per week) | **Duration**<br>(hours per HD session) | **Study Participants**<br>N (%) |
| --- | --- | --- |
| 3 | 3.5 | 14 (10.0) |
| 3 | 3.0 | 13 (9.3) |
| 3 | 2.5 | 1 (0.7) |
| 2 | 4.0 | 5 (3.6) |
| 2 | 3.5 | 4 (2.9) |
| 2 | 3.0 | 4 (2.9) |

Medication burden, as defined by the number of oral, parenteral, and total pills and injections per week, was significantly higher for participants prescribed 720 minutes or more of HD per week than participants prescribed less than 720 minutes of HD per week [Median (IQR): 140.0 (105.0 – 175.0) vs. 112.0 (80.5 – 150.5); p = 0.0011], however, there was no significant difference in the total number of different medications [Median (IQR): 11 (9 – 14) vs. 10 (8 – 13); p = 0.1496].

Despite the differences in frequency and duration of HD, there was no statistically significant difference in median (IQR) ultrafiltration rate (UFR) per session between the two groups [9.3 (7.2 – 11.2) vs. 7.7 (5.7 – 12.0); p = 0.22]. There was also no statistically significant difference in the average (SD) single pool Kt/V Diascan per session between the two groups [1.2 (0.2) vs. 1.2 (0.2); p = 0.3870] (Supplementary Figure 2).

For the KDQOL-36 ^TM^ questionnaire scores, there was no statistically significant difference in 4 of the 5 scales for participants between groups. The participants prescribed less than 720 minutes of HD had statistically significant higher median (IQR) quality of life scores for the ‘Symptoms & Problems List’ scale than the participants prescribed 720 or more minutes of HD [79.2 (70.8 – 88.5) vs. 70.8 (62.5 – 81.3); p = 0.0022).

There was no significant difference in albumin, phosphate, nor calcium laboratory values between groups. The median (IQR) parathyroid hormone laboratory value of participants prescribed 720 minutes or more of HD per week was statistically significantly higher than participants prescribed less HD per week [55.0 (36.0 – 72.9) vs. 40.8 (30.5 – 61.3), p = 0.0396].

In a period of two years, one year before and one year after the date of consent, patient participants who were prescribed less than 720 minutes per week were less likely to visit the emergency department (IRR 0.52, 95% CI 0.33-0.81). However, there was no significant difference between the HD prescription groups and the number of days hospitalized (IRR 0.91, 95% CI 0.44-1.89).

Within one year from the date of consent, 11 of the 41 (26.8%) of the patient participants prescribed less than 720 minutes of HD died, 4 of the 11 (36.4%) of whom opted to withdraw from HD. During that same period, 14 of the 99 (14.1%) of the patient participants prescribed 720 minutes or more of HD died, 3 of the 14 (21.4%) of whom chose to withdraw from HD. However, odds of death did not achieve statistical significance in the unadjusted model (OR 2.23, 95% CI 0.91-5.44) or when adjusted for age and comorbidity (OR 1.62, 95% CI 0.59- 4.49).

## Discussion

In this observational study, patients receiving less than 720 minutes of HD were older and had a lower BMI than patients receiving 720 or more minutes of HD. These characteristics are some of the factors that may have contributed to the shared decision between individual patients and their nephrologist at our regional institution to implement a strategy of prescribing less HD. With advancing age, patients receiving HD often experience metabolic changes, including a reduction in metabolic rate and dietary intake (resulting in lower uremic toxin production) as well as decreased interdialytic weight gain.^24^ Both of these age-related changes support the theory that HD prescriptions may be decreased in these individuals who may not require intensive solute and volume removal. This interpretation is supported by our finding that the median UFR was similar between groups. If interdialytic weight gain had been comparable between groups, the less than 720 minutes of HD group would be expected to require higher UFR to achieve adequate fluid removal. Higher UFR has been associated with an increased risk of intradialytic hypotension, which may negatively affect quality of life due to symptoms of hypotension, as well as increased overall mortality.^25^ Thus, when weighing whether a reduced HD dose is a safe option, lower interdialytic weight gain (such that UFR remains within an acceptable range) should be carefully considered to minimize the risk of symptomatic hypotension, hypervolemia, or emergency department visits for rescue dialysis.

Medication burden has previously been reported to adversely affect quality of life in patients receiving HD. Numerous medications are commonly prescribed in this population, with some patients taking more than 15 pills per day.^26^ Frameworks for end-of-life care in dialysis emphasize deprescribing, allowing for permissive hypertension, less stringent phosphocalcic targets, and diet liberalization in order to prioritize symptom control and reduce treatment burden.^13^ In our study, medication burden was significantly lower in the less than 720 minutes of HD group, a finding that is consistent with principles commonly applied in end-of-life dialysis care.^27^

Despite a lower medication burden, the patient participants in the less than 720 minutes of HD group had lower parathyroid hormone (PTH) values. This finding might reflect baseline differences between groups (specifically that patients in the less than 720 minutes of HD group were older and had a lower BMI) rather than an effect of the HD prescription itself. PTH values have previously been shown to decline with age in patients receiving dialysis^28^ and to increase with higher BMI.^29^

A potential concern with reduced dialysis frequency is the need for rescue HD due to metabolic derangements or hypervolemia. At our centre, patients typically access rescue dialysis through the emergency department. Interestingly, patients in the less than 720 minutes of HD group were less likely to present to the emergency department than patients receiving 720 or more minutes of dialysis.

However, this finding may reflect differences in baseline characteristics between groups, particularly age. In a Canadian study, patients receiving HD aged 18–44 years were more likely to present to the emergency department than older patients.^30^

In addition to the observation of emergency visits, we also quantified hospitalizations and assessed patient-reported quality of life with the KDQOL-36^TM^ questionnaire. Together, these three outcome variables help to show a pattern of important patient outcomes about overall quality of life. In this study, we did not observe any difference in the number of days hospitalized between the two groups. There was no statistical difference in quality of life in 4 of the 5 scales of the KDQOL-36^TM^ instrument. Similarly, other studies have shown that there is no difference in quality of life between participants prescribed thrice weekly versus twice weekly HD.^31^ Based on prior analyses, the statistically significant higher median quality of life score for the ‘Symptoms & Problems List’ scale that we found for the participants prescribed less than 720 minutes of HD is also considered clinically significant.^32^ These patient-reported quality of life results are specific to this study, so further studies are needed to better understand quality of life for patients prescribed a reduced HD dose.

Although we did not find any statistically significant difference in mortality between groups, there was a trend toward increased mortality in the less than 720 minutes of HD group, even after adjustment for age and comorbidity score. This partially unaccounted baseline difference between groups (e.g., patients who have life-limiting diagnoses or poor expected longevity may have been offered a dialysis prescription aligning with palliative goals of care). These findings underscore the importance of individualized HD prescribing, guided by patient-specific treatment goals and shared decision-making.

Our findings are similar to some findings in other small studies such as the DLITE pilot randomized clinical trial.^33^ This Canadian study enrolled 40 patients aged >70 years who were randomized to twice-weekly HD or thrice-weekly HD and followed for nine months. No difference in serious adverse events was observed, although there was a modest increase in the proportion of patients in the twice weekly arm leaving dialysis one kilogram or more above their prescribed weight. This suggests that less frequent HD sessions could be a safe and feasible HD prescribing option for older patients with palliative care treatment goals.

### Study Limitations

Our patient participant enrollment was limited by the single centre nature of this study. Increased statistical power with a higher number of participants may have identified significant trends in mortality. We did not include a frailty assessment of the study participants. Frailty status has shown to be an important factor in overall dialysis mortality^34^ and should be considered for individualized HD prescribing, particularly for elderly patients. Our study was observational, and we did not account for how long patients had been prescribed less than 720 minutes of HD. Patients may have had HD prescription changes within the year from the date of consent and after the data had been collected.

Regarding the patient quality of life assessment, we did not account for the variety of social determinants of health, specifically patient annual income. We chose to use the KDQOL-36^TM^ questionnaire that does not include a question about annual income. The patient medical chart and the provincial health databases were used in this study for data collection, so there was no source document to verify the validity of this patient-reported response given that income is not included in these databases. Further studies are required to assess patient quality of life on reduced HD dose in the context of annual income and the other social determinants of health.

## Conclusions

This study describes the current HD prescribing patterns at our central nephrology clinic and four satellite sites in New Brunswick, Canada. To our knowledge, there are no large, cluster randomized trials that assess the safety and feasibility of varying frequency and duration of HD prescriptions. Our study is important as it may help identify patients who are more likely to benefit from HD dose reduction, such as older individuals with lower interdialytic weight gain. Ultimately, shared decision making with the patient in consideration of treatment goals should direct HD prescribing. When prolonged longevity is not a treatment goal, less dialysis could be considered for patients with palliative care treatment goals which would allow for more comfort and personal time. As a reduced HD prescription has been shown to be appropriate with patients who start dialysis with some residual kidney function,^35,36^ conversely, a reduced HD prescription may be appropriate for older patients for whom the longevity benefit from receiving at least 720 minutes of weekly HD may not be appropriate given patient-specific quality of life values and the inherent higher mortality of this older population.^37^ Overall, a reduction in frequency and duration of HD sessions could be a key factor in end-of-life discussions and planning.

## Supporting information

STROBE Checklist_Cohort

## Data Availability

The authors are willing to share the de-identified data used in the analyses for this study, upon reasonable request.

## Acknowledgements

The authors gratefully acknowledge the collegial collaboration of the nursing teams and the clinic staff of the Horizon Health Network central hemodialysis centre and the four satellite sites for their assistance in identifying potentially eligible study patient participants and for helping to facilitate the informed consent study discussions for the participants and the clinical research manager. The authors also gratefully acknowledge the patients who volunteered to participate in the study and thank them for their interest and their time.

## Statements and Declarations

### Ethical Considerations

This study was reviewed and approved by the Horizon Health Network Research Ethics Board (RS#: 2023-3305).

### Consent to Participate

All patients who volunteered to participate in this study provided written informed consent in either English or French. The original signed and dated informed consent form was saved in the study files and one copy was provided to the participant and one copy was filed in the participant medical chart. The informed consent form for both languages was reviewed and approved by the Horizon Health Network Research Ethics Board prior to the start of the study.

### Consent for Publication

Not applicable.

### Declaration of Conflicting Interest

The authors declared no potential conflicts of interest with respect to the research, authorship, and/or publication of this article.

## Funding Statement

This study was supported by funding, in part, by a Clinical Scholarship awarded to MGM from the New Brunswick Health Research Foundation (currently named Research NB) and by funding from the Saint John Regional Hospital Foundation.

## Notes

### Competing Interest Statement

The authors have declared no competing interest.

