## Supplementary figures and images for "Hemodialysis Prescribing Patterns of Hospital & Satellite Centres: An Institution-Wide Observational Study"

### Supplemental Figure 1

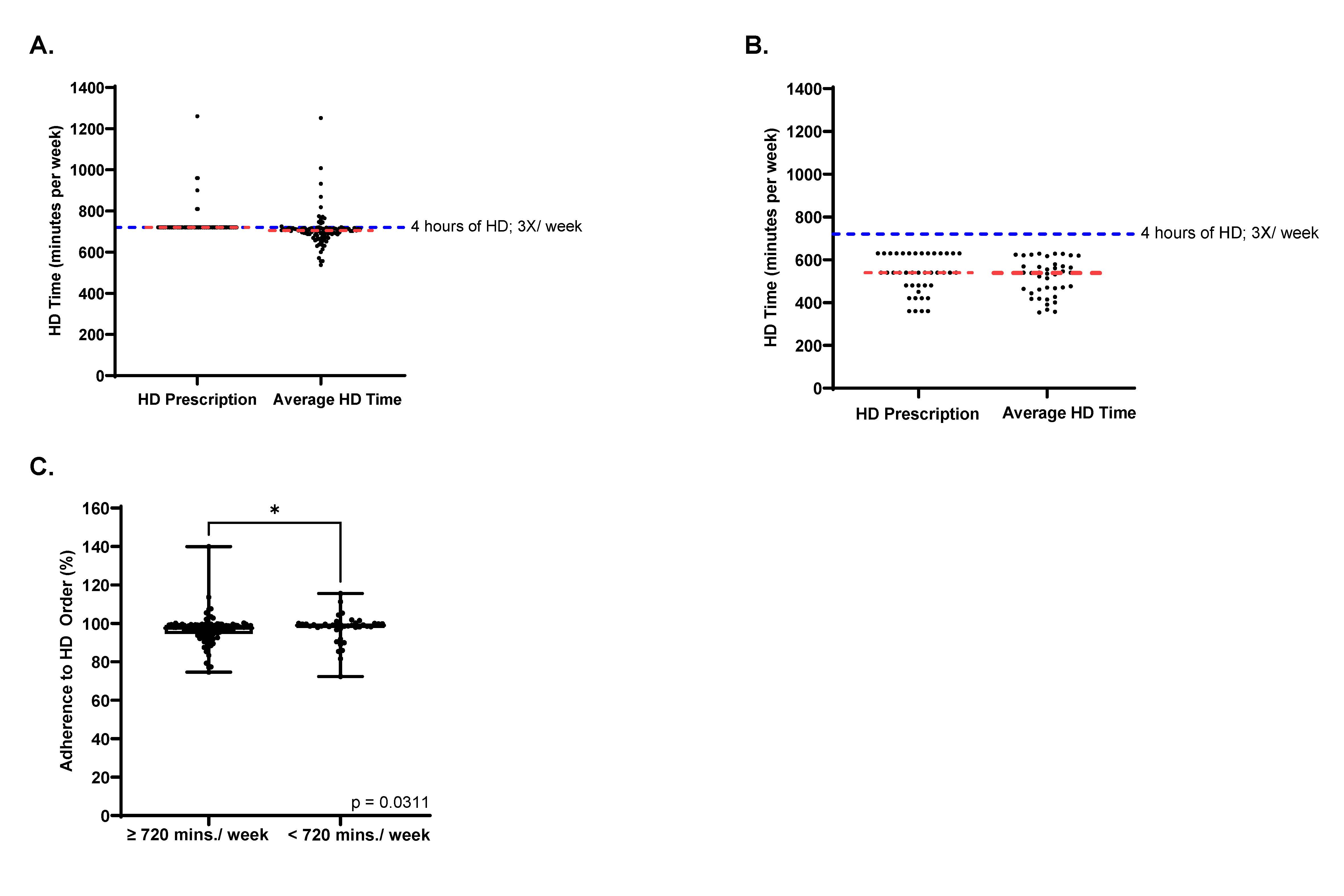

### Supplemental Figure 2

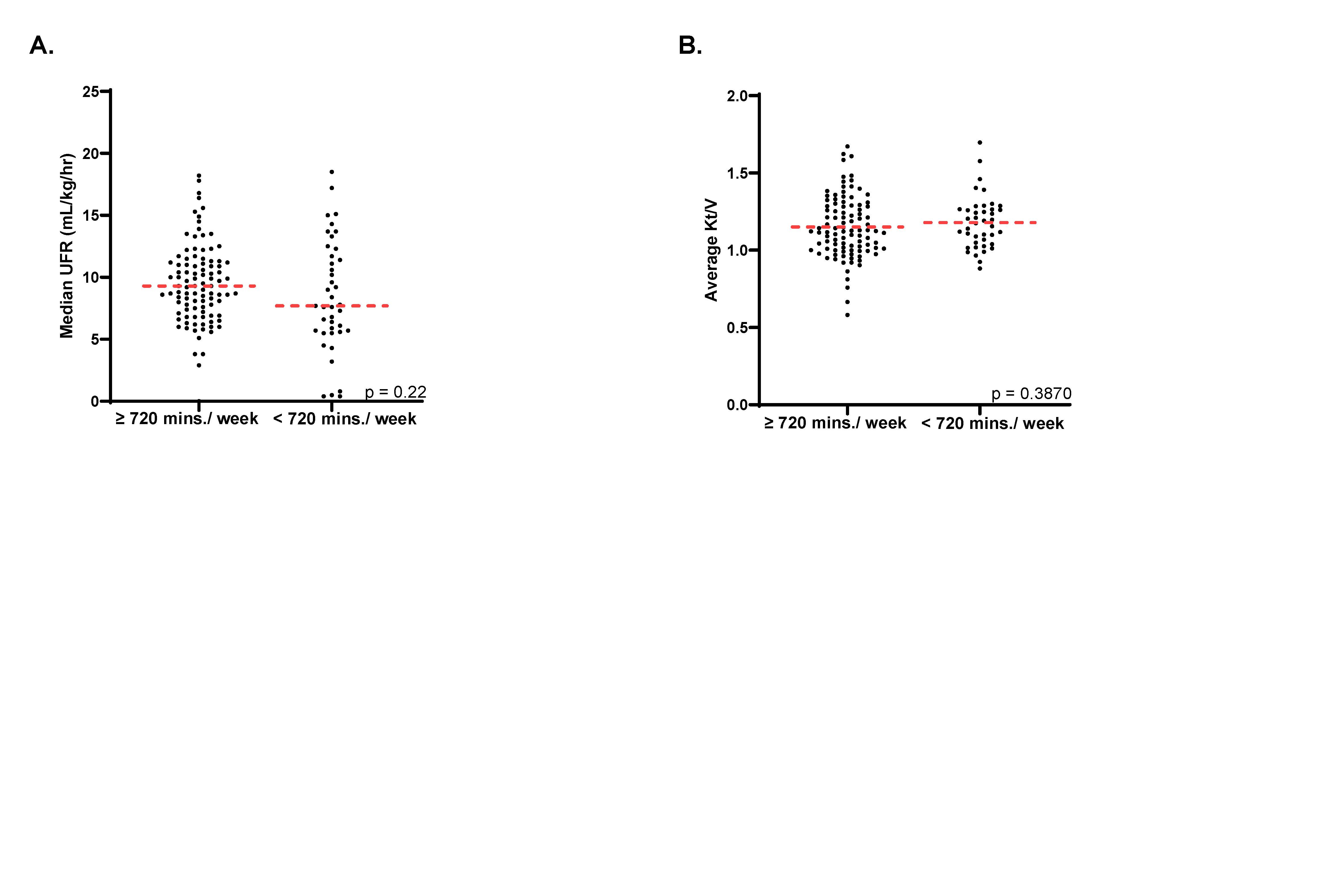
