## Supplemental Figure Legends for "Hemodialysis Prescribing Patterns of Hospital & Satellite Centres: An Institution-Wide Observational Study"

**Supplementary Figure Legends**

**Supplementary Figure 1:** **Study Participant Adherence to Hemodialysis Prescriptions**

(A) The study patient participants observed with a hemodialysis prescription of 720 minutes or more per week versus the average hemodialysis time for 4 weeks (minutes per week), and (B) the study patient participants observed with a hemodialysis prescription of less than 720 minutes per week versus the average hemodialysis time for 4 weeks (minutes per week). (C) The percentage of adherence for 4 weeks to the observed hemodialysis prescription by the study patient participants, respectively.

**Supplementary Figure 2:** **Ultrafiltration & Kt/V Diascan Data**

(A) The median (IQR) ultrafiltration rate (UFR) of the respective study patient participants’ hemodialysis sessions for 4 weeks grouped by hemodialysis prescription of 720 minutes or more per week versus less than 720 minutes per week [9.3 (7.2 – 11.2) vs. 7.7 (5.7 – 12.0), p = 0.22]. (B) The mean (SD) urea clearance (Kt/V Diascan) of the respective study patient participants’ hemodialysis sessions for 4 weeks grouped by hemodialysis prescription of 720 minutes or more per week versus less than 720 minutes per week [1.2 (0.2) vs. 1.2 (0.2), p = 0.3870].
